# Analysis of the malaria profiles in high-risk incidence municipalities in the Brazilian Amazon using Principal Component Analysis in the period 2011-2013 and 2017-2019

**DOI:** 10.1101/2023.07.17.23292751

**Authors:** Natália Martins Arruda, Vinícius de Souza Maia, Bianca Cechetto Carlos, Carlos Eduardo Beluzo, Luciana Correia Alves

## Abstract

**Background:** Malaria still is one of the most relevant infectious diseases in Brazil with 184,869 cases in 2019 and 62.8% of these cases occurred in only 6.2% of municipalities that had high transmission. The incidence of malaria is influenced by environmental, socioeconomic, demographic, and structural factors (organization and effectiveness of health services, land use, and infrastructure).

**Methods:** We use data from the Malaria Epidemiological Surveillance System aggregated by period and municipality, maintained by the Ministry of Health, from 2007 to 2019, and combine it with data from the National Institute for Space Research’s – *Satellite Monitoring of Brazilian Amazon Forest Deforestation Project*, 2010 Demographic Census Microdata and Registry of Health Establishments to analyze socioeconomic, demographic, environmental and health factors in two distinct periods. We use the Principal Component Analysis algorithm to create different principal components characteristics patterns in high-risk municipalities.

**Results:** The Principal Component Analysis allowed the creation of three profiles of high-risk municipalities combining the contribution of different demographic, socioeconomic, environmental, and health characteristics with the contribution of the municipalities in each principal component. The first group with a vulnerability profile, that is, high-risk municipalities with cases more associated with vulnerability characteristics, the second group is one with à profile of occupation and working age, of high-risk municipalities with cases more related to socioeconomic and demographic variables linked with specific occupations and a working-age population and the third group with municipalities that has a mixed profile associated with different characteristics related to of *P. falciparum* in contrast of *P. vivax* cases, such as the municipality of Rio Preto da Eva, which for the cases of *P. vivax*, may be more related to children under 5 years and differently, *P. falciparum* in this municipality could be more related to gold panning, deforestation and the presence of large bodies of water.

**Conclusions:** There are particularities in socioeconomic, demographic, environmental, and health characteristics for infection by both types of *Plasmodium* and high-risk municipalities that become necessary to understand the differences in the profile of the population affected by malaria together with the environment in which they live, the weather, forest, hydrography and health conditions to assess the structure in these regions, in addition to analyzing the cases of *P. falciparum* separately from the cases of *P. vivax*, showing us that understand the socio-environmental determinants at the local level is essential for the success of malaria prevention and control strategies.

**Author Summary:** Luciana C. Alves is an Associate Professor at the Institute of Philosophy and Human Sciences (IFCH) at the University of Campinas (Unicamp) and research scientist at the Population Studies Center ‘Elza Berquó’ (Nepo) and Chair of the Department of Demography at the University of Campinas. Natália M. Arruda is a doctoral student in Epidemiology at the National School of Public Health/Fiocruz. Vinícius S. Maia is a doctoral student at the Centre for Economic Demography at Lund University. Bianca C. Carlos is an assistant researcher at the Population Studies Center ‘Elza Berquó’ (Nepo)/Unicamp. Carlos E. Beluzo is a Professor at the Federal Institute of São Paulo and a doctoral student at the Institute of Philosophy and Human Sciences (IFCH) at the University of Campinas (Unicamp).

## Introduction

Around the world, there were 229 million malaria cases in 2019 in 87 countries. The malaria incidence reduced from 80 cases per 1,000 population at risk in 2000 to 57 cases in 2019. In the Americas, malaria cases have reduced by 40% (from 1.5 million to 0.9 million), in recent years. This progress has been hampered by the major increase of cases in Venezuela, from 35,500 cases in 2000 to 467,000 in 2019. Brazil, Colombia, and Venezuela express over 86% of all cases in the region [1].

Worldwide, *Plasmodium falciparum* malaria is more prevalent than *Plasmodium vivax*. In Brazil, the opposite is true and *P. vivax* cases correspond to 84% of total reported malaria [2]. Despite *P. falciparum* malaria causes higher levels of morbidity and mortality than *P. vivax*, the last one is also very important due to the peculiarities that make it harder to control, as the high number of asymptomatic individuals, undetectable parasitemia, parasite latent stages (hypnozoites), early production of gametocytes and suspicions of resistance to antimalarial drugs [3,4].

Between the period 2011 to 2019, Brazil went from 310,450 positive cases of malaria registered per year to 184,869, a reduction of approximately 60%. As countries accelerate towards malaria elimination, reduction in malaria transmission is highly variable leading to significant heterogeneities in residual transmission across their territories [5,6,7,8].

The Legal Amazon (LA) is an endemic malaria region, consisting of 772 municipalities in the states of Acre (AC), Amapá (AP), Amazonas (AM), Pará (PA), Rondônia (RO) and Roraima (RR), Maranhão (MA), Mato Grosso (MT) and Tocantins (TO). Incidence of malaria varies among municipalities and is measured by the Annual Parasite Incidence (API). Municipalities are classified as having a low risk of incidence when the number of cases is less than 10 per thousand inhabitants, medium risk when it is between 10 and 50 cases per thousand inhabitants, and high risk when it has more than 50 cases. In 2019, 6.2% of municipalities had high transmission and included 62.8% of the total cases.

Although environmental and social conditions are important for malaria-endemic levels, factors such as access and quality of health services act on the dynamics of the disease [9,10]. The interaction of environmental, health and socioeconomic factors favors variations in notifications, many of which are associated with the flaws in epidemiological surveillance actions, responsible for delays in diagnosis, treatment of the disease, and conditions of the vulnerability of the population [11,12].

According to Confalonieri [13], the environmental and social characteristics of the Brazilian Amazon are relevant to the determination of epidemiological patterns. The region’s geographic and ecological characteristics also substantially determine potential habitats for vector reproduction [14]. The incidence of malaria in municipalities is influenced by environmental, socioeconomic, demographic, and structural factors (organization and effectiveness of health services, land use, and infrastructure) [8].

Given the heterogeneity of malaria cases across regions, where there is an epidemiological transition towards elimination in some, but in others, it has remained an endemic disease with sustained transmission, it has become necessary to understand on the aggregate level, the main factors related to the municipalities with the lowest and highest risk of malaria incidence [15,7,5].

Understanding all the factors associated with malaria among high-risk municipalities of Brazil is necessary to promote better malaria control and more effective surveillance actions at a local level. Our choice of period of analysis was the result of our understanding that from 2014 to 2016, the reduction in the number of malaria cases was extraordinary. As a result, we have attempted to compare periods where conditions were similar.

For the aforementioned reasons, the objective of this paper is to analyze the high incidence risk profiles in the municipalities in the period 2011-2013 and 2017-2019, and for this analysis, the Principal Component Analysis (PCA) algorithm was used. Principal Component Analysis (PCA) algorithm is a method used to assist in understanding the most significant relationships between variables, making it possible to analyze how each chosen variable is associated with each component to identify these relationships and used as an exploratory tool [16,17].

## Methods

Data were obtained from the Malaria Epidemiological Surveillance System (SIVEP-Malaria) for the 2011 to 2019 period. This system records malaria cases in the Legal Amazon region, and each malaria-diagnosed patient file is filled with over 40 demographic, socioeconomic, epidemiologic, and health data including service, patient, and disease variables.

The environmental variables – Hydrography, Increment, Cloud, Deforested – were taken from the National Institute for Space Research’s (INPE) *Satellite Monitoring of Brazilian Amazon Forest Deforestation Project* (PRODES). The variable of crude migration was calculated from the 2010 Demographic Census Microdata conducted by the Brazilian Institute of Geography and Statistics (IBGE).

The number of level general care establishments was taken from the Registry of Health Establishments (CNES) which has information on healthcare infrastructure and services, type of care offered, specialized services, number of available hospital beds, and existing capacity, to assist in health planning.

Only positive cases were used, and grouped by the municipality of infection and year of registration, counting the number of occurrences of each variable used in each category.

The results were divided between the periods 2011-2013 and 2017-2019, by high-risk municipality through the API, by species of *Plasmodium* (*P. vivax* and *P. falciparum*), and by a group of variables (Table 1): Environmental and Health; Demographic and Socioeconomic Variables.

**Table 1:**
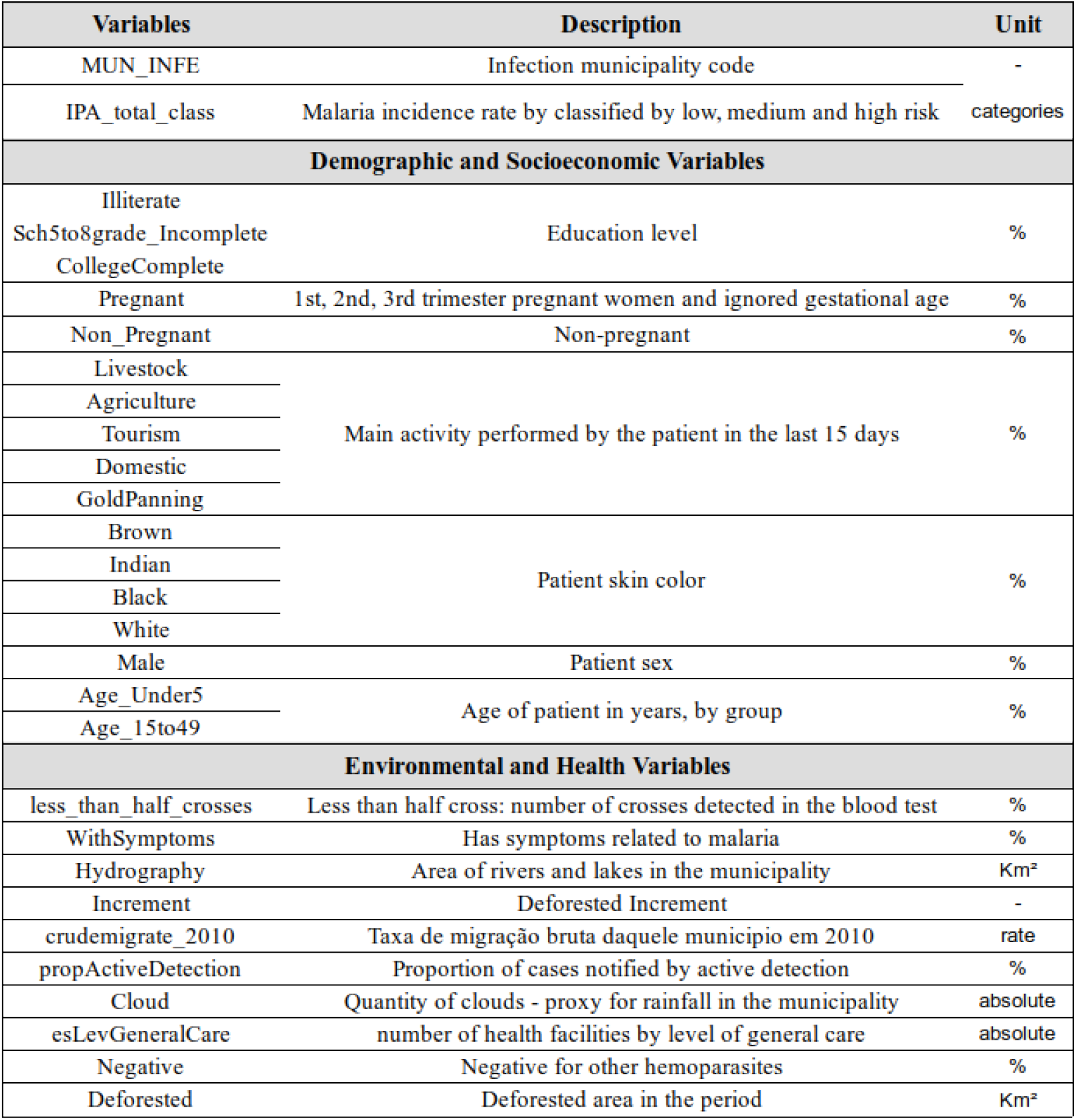
Variables used on the PCAs. Source: SIVEP – Malaria, 2011-2019.

Principal component analysis (PCA) is a method used to show strong patterns between variables by reducing the number of characteristics when building the principal components, sometimes referred to as dimensionality reduction. The principal components make it possible to reduce a set of correlated variables into a smaller number of representative variables that collectively explain most of the variability in the original set [16,17]. For the best performance of the PCA, the data were scaled and centered.

In addition to generating principal components, the method is highly interpretable, allowing the analyst to understand which variables were most strongly associated with each component as well as which data points had a greater influence on the characterization of a given component. Each component is a linear combination of the variables and the coefficient and each variable is called a Loading (factor weights) and the contribution of the municipalities is called Score. The loading helps us understand which variables are most important for each major component [16,17]. Where many variables correlate with one another, they will all contribute strongly to the same principal component.

We have also included a descriptive map of municipalities by the level of risk for the two periods analyzed, to familiarize the reader with the spatial distribution of this risk across the Brazilian Amazon.

The PCA algorithm used was from the *prcomp* package and the models were executed in R version 3.6.3.

## Ethics Statement

This work uses data in the public domain, made available upon request for access to information to the Brazilian Ministry of Health, in accordance with Law No. 12,527, of November 18, 2011 (L12527). The data is de-identified, and the results present aggregated information, therefore, this work is exempt from being evaluated by the Ethics and Research Committee, according to Resolution No. 510, of April 7, 2016, of the National Health Council (Resolution CNS/MS 510/16).

## Results

**F**igure 1 shows the risk of malaria transmission by a municipality in the two periods. The municipalities that went from medium risk to very high risk in the two analyzed periods deserve special attention: Santa Isabel do Rio Negro (state of Amazonas), Alto Alegre, Iracema, Pacaraima, and Rorainópolis (all in the state of Roraima). When we look at the 46 municipalities with high or very high risk in 2017-2019, 41% are municipalities in the state of Amazonas and 24% are in Roraima. In comparison, of the 276 low-risk municipalities, 36% are in the state of Pará, only 7% are in the state of Amazonas and 1% are in the state of Roraima. That is, there is a greater concentration of high and very high-risk municipalities in the state of Amazonas and Roraima and a few municipalities in a low-risk situation in these states.

**Figure 1.**
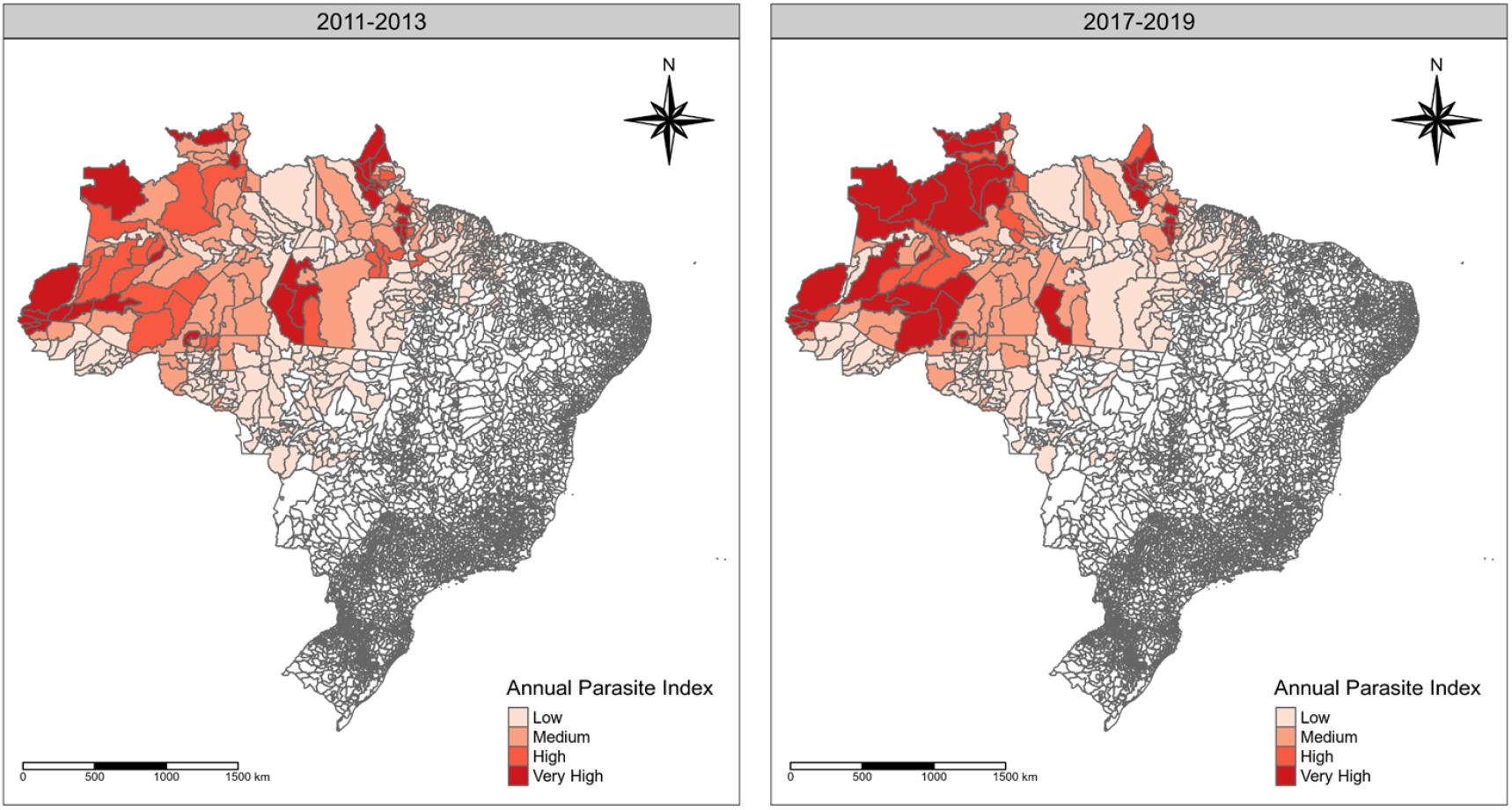
Risk of malaria transmission in Brazil, in the period 2011-2013 and 2017-2019. Source: SIVEP – Malaria, 2011-2013; 2017-2019.

Only 5 municipalities managed to change from high incidence in the period 2011-2013 to low incidence in the period of 2017-2019: São Paulo de Olivença (AM), Tabatinga (AM), Anapu (PA), Goianésia do Pará (PA) and Senador José Porfírio (PA).

There are 48 high-risk municipalities with positive cases for *P. vivax*. Regarding positive cases for *P. falciparum*, we have 48 high-risk municipalities in the first period. Table 2 shows the first 4 main components that represent the highest percentage of variance from the PCA in the period 2011-2013 and 2017-2019, and also facilitates the comparison between *P. vivax* and *P. falciparum* cases, by groups of variables (demographic and socioeconomic; health and environmental).

**Table 2:** Percentage of total variance explained for the four main components selected, 2011 - 2013 and 2017-2019. Source: SIVEP – Malaria, 2011-2013; 2017-2019. *sum of the percentage of the first four components of which one of the models.

| Period 2011 to 2013 |  |  |  |  |  |
| --- | --- | --- | --- | --- | --- |
| Principal Components | *P. vivax* (n=48) |  | Principal Components | *P. falciparum* (n=48) |  |
|  | Demographic and Socioeconomic (%) | Environmental and Health (%) |  | Demographic and Socioeconomic (%) | Environmental and Health (%) |
| PC1 | 30.79 | 31.81 | PC1 | 30.52 | 31.13 |
| PC2 | 12.92 | 15.14 | PC2 | 13.97 | 15.86 |
| PC3 | 11.34 | 15.02 | PC3 | 10.14 | 13.58 |
| PC4 | 10.12 | 10.82 | PC4 | 8.81 | 10.66 |
| **Total Variance\*** | **65.17** | **72.79** | **Total Variance\*** | **63.44** | **71.23** |
| Period 2017 to 2019 |  |  |  |  |  |
| Principal Components | *P. vivax* (n = 48) |  | Principal Components | *P. falciparum* (n = 45) |  |
|  | Demographic and Socioeconomic (%) | Environmental and Health (%) |  | Demographic and Socioeconomic (%) | Environmental and Health (%) |
| PC1 | 36.04 | 27.08 | PC1 | 24.20 | 26.91 |
| PC2 | 14.29 | 18.25 | PC2 | 17.42 | 17.61 |
| PC3 | 10.51 | 15.88 | PC3 | 10.79 | 15.27 |
| PC4 | 9.26 | 10.36 | PC4 | 7.72 | 10.73 |
| **Total Variance\*** | **70.1** | **71.57** | **Total Variance\*** | **60.13** | **70.52** |

There isn’t an important difference in the 2011-2013 period between *P. vivax* and *P. falciparum* concerning the explained variance on the PCA models. In the period 2017-2019, for cases of *P. vivax*, the first 4 main components explain 70.10% in the group of demographic and socioeconomic variables and 71.57% in the group of environmental and health variables. For *P. falciparum* cases, it explains 60.13% and 70.52%, respectively.

### Principal Components Analysis in High-Risk Municipalities

Figure 2 shows the loadings greater than 0.25 in absolute values for the first four principal components of all PCA models.

**Figure 2:**
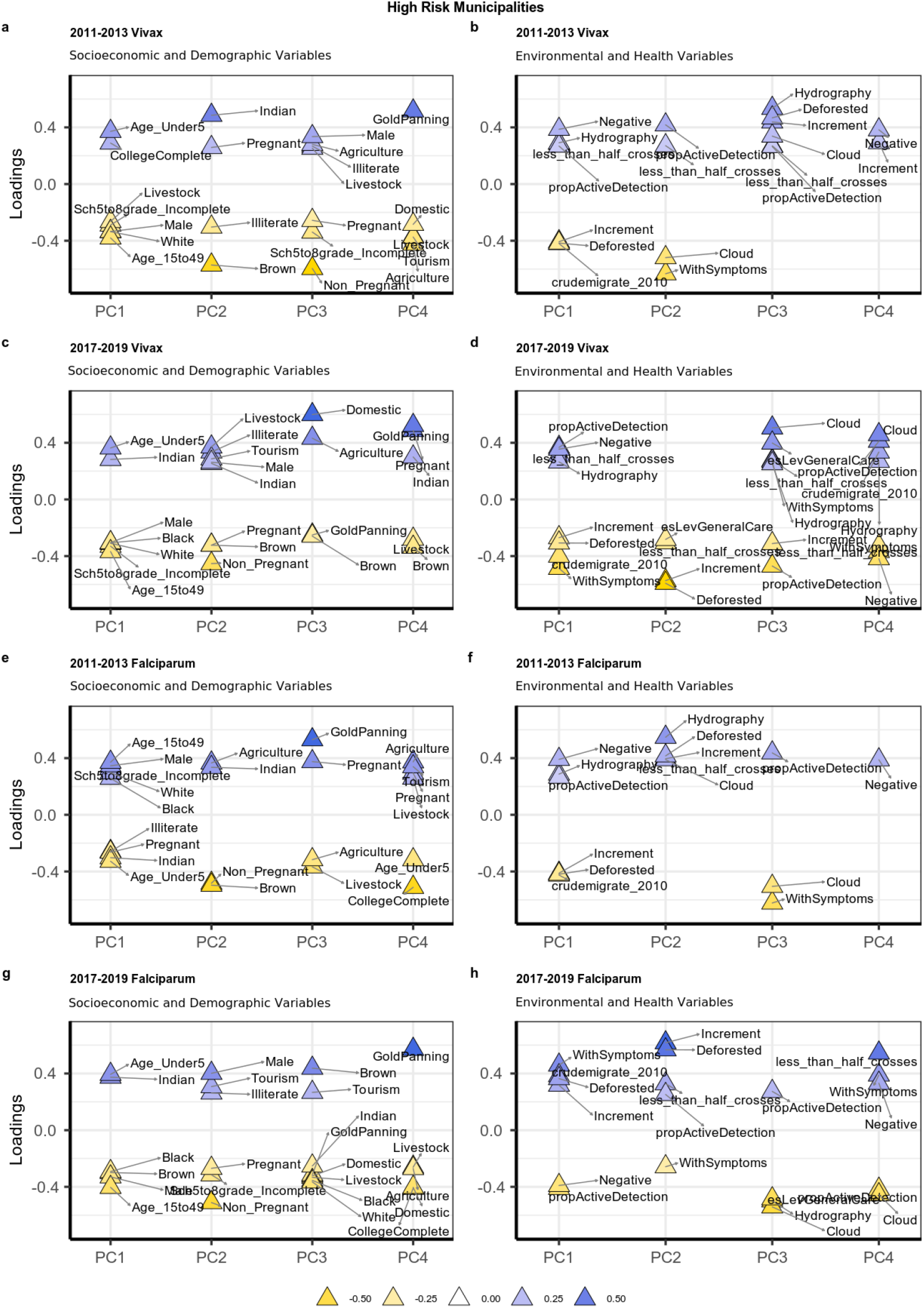
Loadings, High-risk municipalities, *P. vivax* and *P. falciparum*, 2011-2013 and 2017-2019 period. Source: SIVEP – Malaria, 2011-2013; 2017-20.

Looking at the positive cases of *P. vivax* for the period 2011-2013, the first principal component was positively correlated with being under 5 years old with complete higher education and negatively correlated with livestock, incomplete elementary school, male, white, and between 15 to 49 years. The second has only two positively correlated variables – being indigenous and pregnant (PC2 – vulnerable subgroups), and negatively correlated with being illiterate and brown. From environmental and health characteristics (Figure 2b) we see a profile of negative from other parasites, hydrography, has less than half crosses on the exam and proportion of active detection (PC1-most of diagnoses characteristics – Figura 2b). It is worth mentioning that the third and fourth main components have only positively correlated variables, such as hydrography, deforested, increment, cloud, less than half crosses, and proportion of active detection.

From the 2017-2019 period (Figure 2c; 2d), the first principal component (Figure 2c) was positively correlated with age under 5 and indigenous; and negatively correlated with male, black, white, incomplete elementary school, and between 15 and 49 years old. The second principal component has livestock occupation, illiteracy, tourism, male and indigenous as positively correlated, and pregnant, not pregnant, and brown negatively correlated. In Figure 2d, from the first component we can see positively correlated: proportion of active detection, negative to other parasites, less than half crosses and hydrography and negatively correlated with increment on deforested area, deforested, crude migration rate and the presence of symptoms. The second principal component has only negatively correlated variables: number of health establishments, less than half crosses on the exam, deforestation, and increment of deforestation in the period. The third component has a direct association with environmental variables (cloud, hydrography, deforested, and increment).

Regarding infection by *P. falciparum*, we see a change in the profile in the two periods. In the first period (Figure 2e), is 15 to 49 years old, male, incomplete elementary school, white and black are positively correlated to the first principal component and negatively correlated with illiterate, pregnant, indigenous, and under five years old. The second PC has an occupation in agriculture and indigenous people as positively correlated, and non-pregnant women and brown negatively. Hydrography and proportion of active detection were positively correlated with the first principal component (Figure 2f).

In the period 2017-2019 instead (Figure 2g), under 5 years old, indigenous are positively correlated, and black, brown, and 15 to 49 years old are negatively correlated. The second principal component has male, tourism, and illiterate as positively correlated and pregnant and not pregnant and elementary education incomplete negatively correlated. When we look at environmental and health variables (Figure 2h), the presence of symptoms, crude migration rate, deforested and Increment were positively correlated and negative for other parasites and the proportion of active detection was negatively correlated.

Figure 3 shows all the high-risk municipalities with positive scores (contribution of the observations – municipalities for every Principal Component) in the recent period of 2017-2019, so from each component we can see the municipalities that most contribute to a specific principal component and suggest that these municipalities have the profiles shown at Figure 2. As we can see, it has some spatial patterns, fundamentally about environmental and health variables.

**Figure 3.**
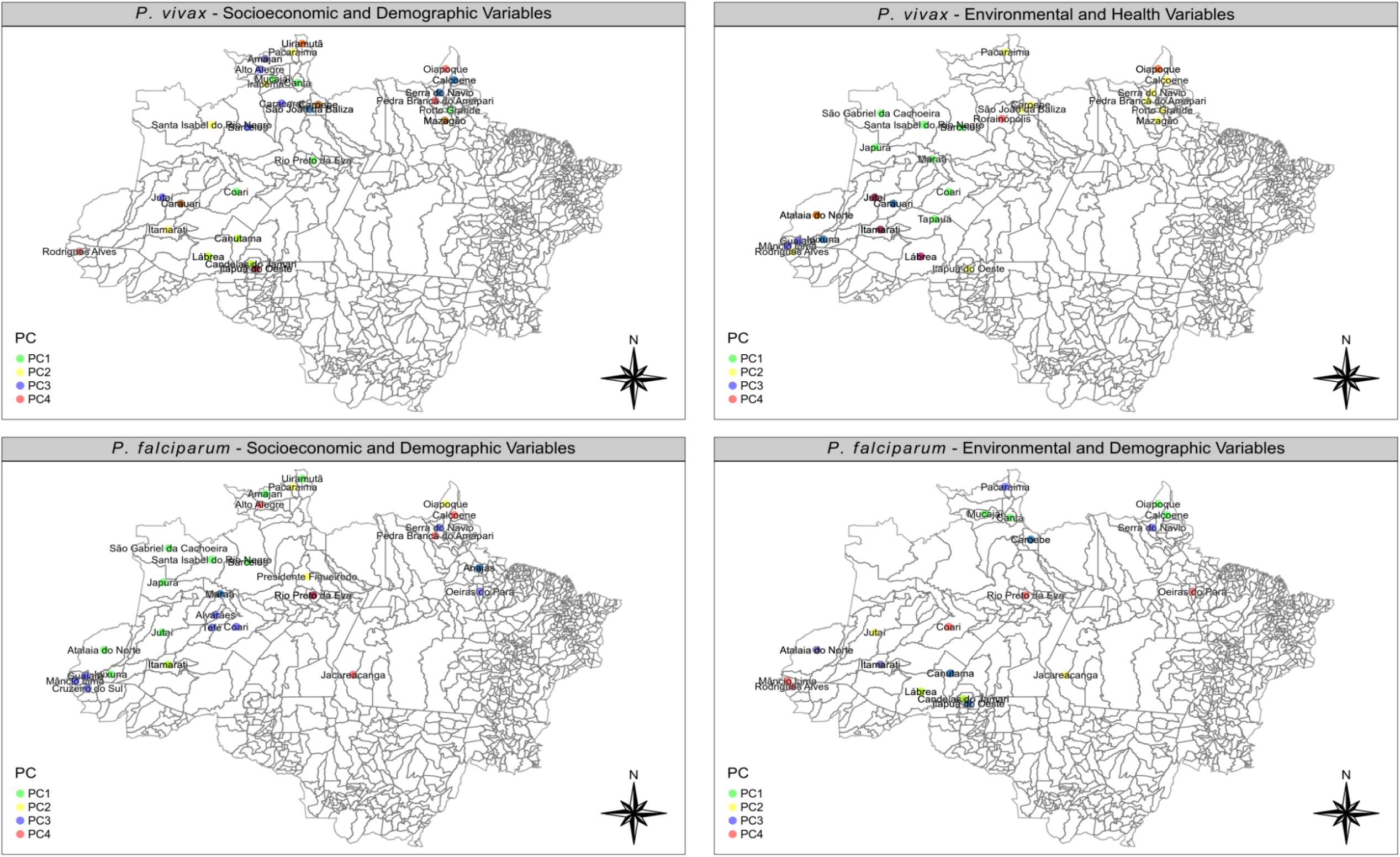
Scores High-risk municipalities, *P. vivax*, and *P. falciparum*, 2017-2019 period.

The municipalities represented by green circles have a profile of cases under 5 years old and indigenous (Figure 3a) for the 2017-2019 period, the group of municipalities with a yellow circle has a profile including livestock, illiterate, tourism, male and indigenous. The PC3 group of municipalities has a profile of Domestic and also Agriculture, and the PC4 group Gold Panning, Pregnant and indigenous.

Relative to Environmental and Health variables (Figure 3b) for *P. vivax*, the first group of municipalities has higher positive loadings for the proportion of active detection cases, negative to other parasites, with parasitemia less than one cross and Hydrography area. The second group (PC2 – yellow circle) has a profile of only negative correlation variables: number of health establishments, deforested area, and increment of deforested area. The third group (PC3 – purple circle) has a high correlation with the number of healthcare establishments, the area covered by water bodies, and parasitemia with less than one cross, and the fourth group (PC4 – orange circle), high loadings of the proportion of active detection and crude migration rate.

Regarding the *P. falciparum* cases and the socioeconomic and demographic profile (Figure 3c), the PC1 group of municipalities with high scores has an age under 5 and an indigenous profile. The second group (PC2) of municipalities has a male, tourism, and illiterate profile. The third group (PC3) of municipalities is associated with a brown and tourism profile and the fourth group (PC4) is associated with a gold panning occupation profile.

Figure 3d shows the municipalities related to the *P. falciparum* cases and Environmental and Health profiles and we can see that the first group of municipalities (PC1) characteristics is associated with crude migration rate, symptoms, deforested area, and increment of deforested area. The second group is also associated with deforested area and increment of deforested area and proportion of active detection and exam with less than one cross (PC2). The third group (PC3) has a profile of proportion of active detection and the fourth group (PC4) has a profile mostly of diagnostic variables: less than one cross, the proportion of active detection, with symptoms and negative to other parasites.

From this assessment, combining the loadings (Figure 2d;e;f;g) with the characteristics and scores (Figure 3) of municipalities we can draw the following profiles when we look at high-risk municipalities in the recent period (2017-2019):

#### Group 1

vulnerability profile, that is, high-risk municipalities with cases more associated with socioeconomic and demographic variables with vulnerability characteristics, such as children and the indigenous population, high proportions of illiteracy, incomplete secondary schooling, and little or no association with environmental or health variables, to the extent that they’re captured in our data.

#### Group 2

profile of occupation and working age, of high-risk municipalities with cases more related to socioeconomic and demographic variables linked with specific occupations and a working-age population, such as men, people aged between 15 and 49 years, with low education and in occupations such as agriculture, livestock and gold panning in addition to environmental characteristics related to deforestation and hydrography.

#### Group 3

profile associated with different characteristics of *P. falciparum* and *P. vivax*, such as the municipality of Rio Preto da Eva, which for the cases of *P. vivax*, may be more related to children under 5 years and differently, *P. falciparum* in this municipality could be more related to gold panning, deforestation and the presence of large bodies of water (high loading in PC4). A few other municipalities appear to follow the same differentiated profile between the two types of *Plasmodium*, such as Coari, Canutama, and São Gabriel da Cachoeira.

It should be noted that the same municipality may have characteristics in more than one group above.

## Discussion

This study analyzed the different high incidence risk profiles in the municipalities in the period in the Brazilian Amazon between 2011-2013 and 2017-2019. It was observed that the risk of having the disease was not uniform, which can be verified by the results of the annual parasite index (API). The results show us that the main components described the majority of variation and explained the varied influence of the original characteristics. These influences, the loadings, show us different characteristics of the municipalities with high API.

There is a greater positive correlation for being under 5 years old, but also with complete higher education, men in the two periods of time, corroborating the finding of the study by Lana [7] which indicated that municipalities with higher API are highly characterized by peri-domestic transmission in which all segments of the population have reported cases and also indicates few changes in the infection profile between the two periods for these high-risk municipalities which is a concern because, unlike low-risk municipalities, they show no improvement trends, maintaining transmission or even increasing. [18], when studying the incidence of malaria and long-term epidemics, showed that municipalities with long-term epidemics were more frequent in the states with the highest incidence.

The study by Lana [7] demonstrates that there are distinct risk profiles between the municipalities and their transmission configurations, for example, notes that in a high-incidence municipality such as Anajás (Pará), a large proportion of cases occur in children. By contrast, in some low-risk municipalities, the majority of cases affect men aged 15 to 65 years old.

We found that municipalities with a high-risk transmission have heterogeneous profiles where, in some municipalities, there will be a greater relationship between the high number of cases with population vulnerability (such as age, indigenous population, education, etc.) and other groups of municipalities, the high-risk incidence could be more related to men in working age and be related to occupation.

The present study shows a predominance of high risk of Falciparum in border regions and mining regions, although other highly vulnerable occupational groups, such as tourism and agriculture, are also observed.

The gold panning occupations are generally located in areas with difficult access, with limited or non-existent health services and, in some cases, they operate illegally, factors that make it difficult to cover timely diagnostic services. In addition, in some states that share an international border, for example, there is evidence of imported cases whose origin is from prospecting in other countries, but which are not registered as such. Another problem of malaria in mining is the high rate of *P. falciparum* (36%) and late and/or inadequate diagnosis and treatment [19].

Figures 2 and 3 show us municipalities with high scores on the groups of socioeconomic and demographic variables but don’t appear on the maps of environmental and health groups. For example, the municipalities of Uiramutã (Roraima) and Rio Preto da Eva (Amazonas) have high scores on the demographic and socioeconomic variables and this could indicate that the high incidence cases of malaria in these municipalities are more strongly associated with population vulnerability than environmental and health variables when it comes to *P. vivax* infections.

Related to Group 1 presented in the previous section, studies show that the lower the years of education the higher the chances to have malaria and this could be related to a lack of opportunity, worse quality of life, and a general context of vulnerability [21,6,22]. Additionally, the high loading of the variable children under 5 years old could suggest that a significant proportion of malaria transmission occurs within households in these municipalities [12]. Children are population groups that have little immunity. Indoor contamination suggests the existence of precarious housing that does not offer protection, increasing contamination.

Group 2 shows us some municipalities with a high association with occupational and environmental characteristics. Corroborating with some studies [23,24,18] that indicate an association between the increase in cases of malaria and gold mining activity, as well as the association between deforestation for agriculture and livestock. Mining contributes to the spread of the disease, as it favors deforestation and the creation of pools of water, an ideal habitat for the reproduction of vectors that transmit malaria.

For instance, Oliveira [24] when analyzing the occurrence of malaria cases in municipalities in Amapá and its relationship with mining activity, found out that in the places where mining projects had been implemented, the number of cases was much higher compared to the places that did not have mining activities. In addition, this profile is more associated with men, which can be explained by the fact that these occupations are generally associated with a male workforce, generally working in areas with poor infrastructure and favorable to the development of disease vectors. Thus, the predominance of cases in male populations in these municipalities in Group 2 is greater due to their higher exposure profile [25,26,27].

The associations between types of economic activity, social mobility, and regionalization of territorial occupation and the incidence of malaria confirmed that malaria infection could be associated with productive activity and labor flow linked to extractive activities and agricultural settlements as found in some principal component results like gold mining, agricultural and livestock occupation [8].

Hydrography and deforestation variables appeared with high loading and positive correlation in high-risk municipalities in Group 2. The high importance of the hydrography variable indicates a relationship between the cases of malaria and the high availability of natural breeding sites for mosquito vectors, since rainfall, river water levels, and climatic factors have been linked with malaria cases [14].

In a prior study, [28] also analyzed the influence of some variables such as temperature, precipitation, and water level on the peaks of malaria cases in four municipalities of Amazonas, and observed a statistically significant correlation between all analyzed variables and causes of disease. This result is explained by the fact that seasonality and the abundance of rainfall cause fluctuations in river water levels and consequent flooding create a favorable environment for mosquito breeding.

Relative to the high loading value in variables such as deforested area (total area deforested in the period) and increment in deforestation (difference in a deforested area in the period), there are some contradictory findings in the literature. While deforestation increased malaria incidence in some studies [29,30,31], forest conservation and high forest cover were associated with a higher incidence in other studies [32,33].

The relationship between deforestation and malaria is the object of some debate since there seems to be no one direction of causality and several concomitant factors may be interacting to determine it. For instance, in the case of “frontier malaria”, the authors identify biological, ecological, and sociodemographic phenomena operating at distinct spatial scales to define it. Here, our inclusion of these variables is an attempt to include an ecological component that contributes to the axis of ecological receptivity of the area to disease vectors, rather than an attempt to summarize the effect of environmental variables on malaria incidence, such as the one attempted [34] for deforestation in several countries in the African continent [35].

Group 3 shows us municipalities with different characteristics between those who are infected by *P. vivax* and those who were infected by *P. falciparum*. [33] found some environmental differences between these two species, for example, *P. falciparum* malaria had a stronger relationship with deforestation than *P. vivax*. In addition, these municipalities show different patterns in socioeconomic and demographic characteristics, such as Rio Preto da Eva (Amazonas), in which transmission by *P. vivax* is related to children under 5 years and the indigenous population, suggesting a more frequent peridomestic infection, while in cases of *P. falciparum* is more related to gold panning.

According to the 2020 Epidemiological Bulletin, in the Amazon region, where 99.9% of malaria cases occur, around 80% of malaria was concentrated in 41 municipalities in 2019, with 16 in Amazonas (39.0%), 8 in Pará (19.5%), 7 in Roraima (17.1%), 4 in Amapá (9.8%), 3 in Acre (7.3%), 2 in Rondônia (4.9%) and 1 in Mato Grosso (2.4%). The National Program to Combat Malaria (PNCM) prioritizes the municipalities that together are responsible for 80% of the country’s autochthonous cases of malaria. The present study uses machine learning algorithms for PCAs to find complex patterns and reinforces priority areas or municipalities previously established by the PNCM, as well as pointing to epidemiological indicators in Brazil.

The use of observational data and some contextual data from other sources is a limitation of this study because the authors didn’t control the study design and quality of data reporting. The inclusion of variables from distinct sources represents an attempt to control for confounding variables in a setting where primary data collection is not feasible.

The objective of this work was to draw profiles associated with the risk of incidence in the municipalities by combining data from the malaria surveillance and control data with environmental and health data from the municipalities.

It is necessary to understand the profile of the population affected by malaria together with the environment in which they live, the weather, forest, hydrography, and health conditions to assess the structure in these regions, in addition to analyzing the cases of *P. falciparum* separately from the cases of *P. vivax*, understanding there are particularities in socioeconomic, demographic, environmental and health characteristics for infection by both types of *Plasmodium*.

Thus, understanding the socio-environmental determinants at the local level is essential for the success of malaria prevention and control strategies.

## Data Availability

This work uses data in the public domain, made available upon request for access to information to the Brazilian Ministry of Health, in accordance with Law No. 12,527, of November 18, 2011 (L12527). The data is de-identified, and the results present aggregated information. Will be available at the acceptance of the paper.

## Acknowledgments

We thank the Bill & Melinda Gates Foundation (Process ID INV 003970), the Brazilian Ministry of Health, the Department of Science and Technology (DECIT), and the National Council for Scientific and Technological Development (CNPq) (Process 443048/2019-3) by financial support.

